# Modified Preparation Method of Ideal Platelet-Rich Fibrin Matrix (PRFM) from Whole Blood

**DOI:** 10.1101/2021.05.20.21257494

**Authors:** Mirta Hediyati Reksodiputro, Alida Roswita Harahap, Lyana Setiawan, Mikhael Yosia

## Abstract

One bioproduct that is widely used in the wound healing process is Platelet Rich Plasma (PRP). PRP is a liquid solution with high autologous platelet concentration, making it a good source of growth factors to accelerate wound healing. Recent development in PRP had created a new product called Platelet Rich Fibrin Matrix (PRFM), which has a denser and more flexible structure. PRFM is the newest generation of platelet concentrate with a fibrin matrix that holds platelet in it. The key concept in creating PRFM from PRP is the addition of CaCl_2_ followed by centrifugation, which converts fibrinogen to fibrin, and the fibrin cross-links to form a matrix that contains viable platelets. There are many commercially available kits to create PRFM, but they are often expensive and uneconomical. This research will test a modified method of making ideal PRFM from PRP without any commercial kits. The modified method will include determining the minimum level of CaCl_2_ used, the type of centrifuge, and the speed and duration of centrifugation. By performing a modified preparation method on five samples of whole blood, it was found that the ideal PRFM could be made by mixing PRP with 25 mM CaCl_2_ 1M and centrifuging it at a speed of 2264 G for 25 minutes at room temperature. The PRP and PRFM platelet counts of this method tend to be lower than the platelet counts found in other studies. Although visually comparable, further study is needed to compare the performance of PRFMs made with this method and PRFMs made with commercial kits.

## 1. Introduction

Otorhinolaryngologist specialists in plastic reconstruction have reported successful use of exogenous growth factors and PRP in clinical settings. Sclafani reported that the release of growth factors in wound healing was primarily carried out by platelets, echoing that PRP plays a vital role in wound healing.^1^ The influence of growth factors on endothelial cells and fibroblasts will increase within seven days after injury and disappear after fourteen days. Other studies had also reported that PRP can increase and accelerate wound healing by 80% in ulcers compared to placebo.^2,3^ In clinical settings, people often use PRP in the form of a solution or gel to facilitate tissue repair. The macroscopic structure of conventional PRP is inadequate because it requires implantation at a specific site, and, likely, the growth factors are not appropriately fixated during surgical procedures.

Platelet Rich Fibrin Matrix (PRFM) is the latest generation of platelet concentrates with simple preparation without biochemical ingredients (bovine thrombin). PRFM is a slow polymerization of fibrin in PRP, resulting in a PRFM structure that resembles natural fibrin.^4^ This specific structure will play an essential role in increasing cell migration, cell proliferation, and cyclic formation. Through this polymerization process, all platelets in PRP will be deposited between the PRFM fibrin fibres. According to that logic, levels of platelet (and subsequently, growth factor) in PRFM are expected to be equivalent to PRP.

Laboratory experiments looking at usage of PRFM in specific medias shows that there was an increase in levels of platelet-derived growth factor (PDGF), vascular endothelial growth factor (VEGF), basic fibroblast growth factor (bFGF), and transforming growth factor beta (TGFβ) on the first day, followed by a gradual decrease on the next day.^1,5^ This characteristic was not observed in PRP since most of the growth factors in PRP were released on the first day of application to the wound. PRFM also has fibrin characteristics with a more natural platelet distribution that mimics the body’s response to injury and a denser and more flexible macroscopic structure.^6,7^ These factors theoretically make PRFM superior to use for wound healing.

Commercial kits for preparing PRFM had been available and widely used in clinical settings. One such example is the FIBRINET tubes which can produce PRFM from whole blood through the addition of CaCl2 and centrifugation at 1100 G for 6 minutes.^5^ However, the use of the aforementioned commercial tools has several drawbacks, including (1) High prices, rendering the use of PRFM in a clinical setting to be economically dubious; (2) concentration of platelet in PRFM is unknown, increasing the possibility of not achieving the creation of ideal PRFM. This study aims to overcome the problems mentioned above and cover the weaknesses of the existing invention by proposing a modified method to produce PRFM.

In order to guarantee the production of ideal PRFM, the modified method will ensure that Platelet Poor Plasma (PPP), a by-product in PRFM production, has a platelet content of 0/μL, representing the fact that all platelets are attached to the PRFM fibrin matrix at the bottom of the tube. Measurement of TGFβ1 will be conducted to reassure that growth factors are still available and viable inside the end product. The modified method will also try to make PRFM production more economical by creating PRFM from PRP through improvised methods without using a commercial PRFM production kit. At the same time, this study will determine the minimum amount of CaCl_2_ needed to produce an ideal PRFM.

This study will carry out trials processing of human’s whole blood, making PRP and adding in 1 M of calcium chloride (CaCl_2_) before centrifuging the solution to form the PRFM. The trials will evaluate whether the modified method can prepare ideal PRFM to accelerate the wound healing process. Improvement in wound healing is essential in reconstructive surgery and will affect the successes of operations both aesthetically and functionally.

## 2. Materials and Methods

This study was conducted in Indonesia and approved by the Ethical Clearance Committee from the Faculty of Medicine, University of Indonesia (202/H2.F1/ETIK/2013). The study’s design and protocol are made and implemented following the Helsinki Declaration. Volunteers recruited from the study were given an explanation, information sheet and signed the informed consent form before participating in the study.

### 2.1. Whole blood collection from participants

Whole blood samples (8 mL per tube) was collected from 5 healthy volunteers. The volunteers are non-smokers, with no history of chronic diseases, and with normal hematologic profile (including normal thrombocyte count, PT/APTT). Blood is collected using the 10 mL vacuum tube with cell selector gel from the PRP kit (RegenKit® A-PRP®, Regen Lab, Le Mont-sur-Lausanne, Switzerland). A small amount of whole blood was also taken to analyse thrombocyte count with the Celtac-a automatic cell counter (Automated Hematology Analyzer MEK-6450, Japan). The kit from RegenLab is designed to produce 4-6 mL of PRP from every tube of blood collected. The RegenKit® vacuum tube was inverted back and forth three times after blood collection is finished.

### 2.2. Preparation of PRP and calculation of thrombocyte count from PRP-PPP (Platelet Poor Plasma)

Blood that had been taken from volunteer were immediately centrifuge using RegenLab PRP-Centri CH-1052 (Regen Lab, Le Mont-sur-Lausanne, Switzerland). The centrifuge from RegenLab was designed to prepare PRP and was programmed to run at 1,500 G for 5 minutes at room temperature. The centrifuge process resulted in three separate layers: a clear yellowish plasma thrombocyte on top (consisting of a PPP layer on top and a PRP layer in the bottom), a cell selector gel layer with leucocyte in the middle, and a red blood cell layer on the bottom. A small amount of PPP and PRP was taken from each specimen to analyse thrombocyte count using Celtac-a automatic cell counter. PRP process is determined to be successful when the thrombocyte count in PPP is 0/μL. The tube was then inverted gently back and forth three times to mix the plasma, thrombocyte, and leucocyte, forming the final PRP. PPP was mixed with PRP to ensure a larger volume of end product used for PRFM preparation. A total of 5 PRP specimens will be produced at the end of this step and will be used to produce PRFM.

### 2.3. Preparation of PRFM and calculation of thrombocyte count from PRFM-PPP

This experiment concentrates on creating PRFM using methods proposed by O’Connel, which is a modified Fibrinet (Cascade Medical Enterprise, Wayne, Ney Jersey) PRFM kit method.^8^ RegenLab PRP-Centri CH-1052 centrifuge was used (Figure 1), as it is the ideal centrifuge to produce PRFM with minimal addition of CaCl_2_. The maximal speed of this centrifuge is 4,500 RPM (2,264 G). Six mL of PRP were taken from 4 different PRP specimens using a micropipette (extra care was given to avoid taking the red blood cell layer) and moved to 4 separate 10 ml cylindrical centrifuge tubes (Pyrex®, Staffordshire, England). Each tube was then added with either 70 mM, 45 mM, 25 mM, or 15 mM of CaCl_2_ 1M. All four tubes were then centrifuged at 2,264 G for 25 minutes at room temperature. The resulting product will consist of two layers, the PRFM and PPP.

**Figure 1.**
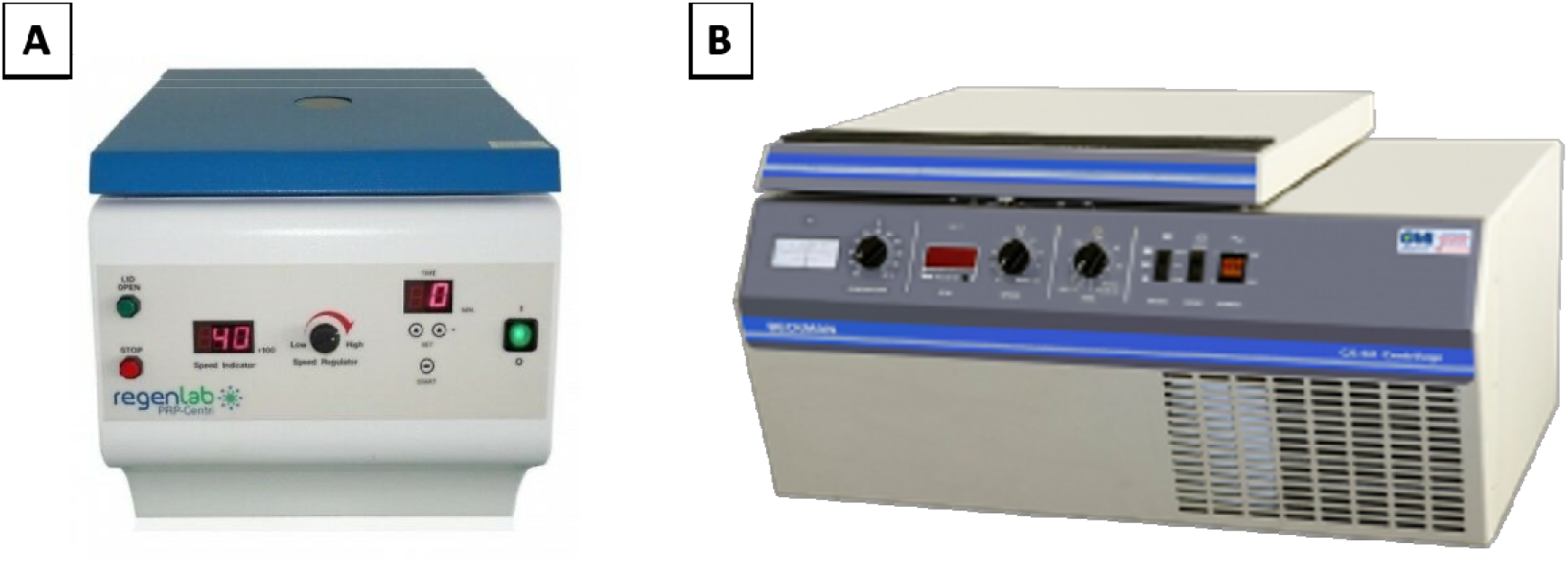
(A) RegenLab PRP-Centri CH-1052 centrifuge used in processing the PRP in cylinder tubes and (B) Beckman CS-6R centrifuge used in processing PRP in a round plastic container.

PPP from all four PRFM specimens with different concentrations of CaCl_2_ was then analyzed using the Celtac-a automatic cell counter. As previously mentioned, an ideal PRFM is the one with a 0/μL thrombocyte count in their PPP. The lowest concentration of CaCl_2_ that produces an ideal PRFM was then determined as the minimum recommended concentration of CaCl_2_.

After the minimum amount of CaCl_2_ needed had been obtained from the previous steps, the 5^th^ PRP specimen was assigned to an alternative PRFM preparation method to produce a coin-shaped PRFM. Six mL from the 5th PRP specimen were taken using a micropipette and put into a round plastic container (diameter 30 mm), followed by addition of the CaCl_2_. The plastic container was then centrifuged with a Beckman CS-6R centrifuge at 3,800 RPM (4,043 G) for 25 minutes (Figure 1). The resulting product will consist of two layers, the PRFM and PPP. The PRFM and PPP was taken for thrombocyte count analysis using the Celtac-a automatic cell counter.

### 2.4. Analysis of PRFM characteristic with Scanning Electronic Microscope (SEM)

The PRFMs are trimmed into equal sizes, washed using phosphate-buffered saline (PBS), and fixated in 2.5% glutaraldehyde in PBS for 1 hour at 4°C. The samples were then washed for the second time with 0.1 M cacodylate buffer (pH 7.3) followed by fixation with 1% osmium tetroxide (OsO4) and 0.1 M cacodylate buffer for 1 hour at room temperature (22 ± 2°C). The samples were then dehydrated in serial ethanol and dried before placed on an aluminium sheet with silver adhesive paint coated with a layer of 4 nM of gold inside an Edward S150B argon atmosphere apparatus (Crawley, West Sussex, UK). The samples were then observed at 0° with SEM Stereoscan 200 (Cambridge, UK) at 20 kV. Three different fields of view were examined for each sample, and the diameter of thrombocyte and size of fibrin fibres were examined.

### 2.5. Analysis of TGFβ1 in PRFM

TGFβ1 in PRFM were analyzed using a TGFβ1 immunoassay kit (Quantikine R&D Systems, Fisher Scientific, New Hampshire, United States). The immunoassay kit works by identifying reactions of growth factors in the sample with TGFβ1 monoclonal antibody in the microtiter plate wells. The bond between TGFβ1 and anti-TGFβ1F was identified via TGFβ1 polyclonal antibody that is labelled with an enzyme. With additional substrates, these reactions will produce a varying intensity of colours identified with spectrophotometry at 450 nm. The intensity of the colour is directly correlated to the concentration of protein analyzed. Compared to a standard solution with a known concentration, the concentration of TGFβ1 in the PRFM sample can be analyzed.

PRFM specimens analyzed were put into the microtiter plate wells coated with TGFβ1 monoclonal antibody and incubated. TGFβ1 in the sample will bind to the antibody inside the well. After washing the well to remove excess substances, the TGFβ1 polyclonal antibody with Horse Radish Peroxidase (HRP) label was added. Second incubation was done, during which the polyclonal antibody will bind to the anti-TGFβ1*TGFβ1 complex that formed in the first incubation forming a sandwich of anti-TGFβ1*TGFβ1*anti-TGFβ1.HRPO. A substrate is added to form blue colour that will change into yellow after a stop solution is added. The intensity of colour will correlate with the amount of TGFβ1 inside the sample.

## 3. Results

### 3.1. Volunteers baseline data and examination of PRP

Blood was drawn from all volunteers and was processed right away using the methods above. All five volunteers (four male and one female, age range 25-45 years) had a normal hematologic profile, no chronic comorbidities, and normal CBC (haemoglobin, hematocrit, thrombocyte, leucocyte, differential counts) examinations. Comparison of the amount of thrombocyte in whole blood, PRP, and PPP can be seen in Table 1.

**Table 1.** Thrombocyte count (per μL) for whole blood, PRP and PPP after preparation of PRP. Note that the thrombocyte count for PPP is 0/μL for all the samples, indicating a successful creation of ideal PRP.

| Sample | Thrombocyte count ($\mu\text{L}$) | | |
| --- | --- | --- | --- |
|  | Whole Blood | PRP | PRP-PPP |
| Subject 1 | 313.000 | 52.000 | 0 |
| Subject 2 | 414.000 | 64.000 | 0 |
| Subject 3 | 205.000 | 66.000 | 0 |
| Subject 4 | 280.000 | 121.000 | 0 |
| Subject 5 | 393.000 | 186.000 | 0 |

### 3.2. Determination of the minimum recommended concentration of CaCl_2_ for ideal PRFM

Four PRP specimen were assigned to different amounts of CaCl_2_ to determine the minimal amount in which ideal PRFM (ones with 0 μL of thrombocyte in its PPP). The schematic shown in Figure 2 shows that 25 mM of CaCl_2_ 1M is the minimum amount needed to produce ideal PRFM. The fifth specimen were used to produce a coin-shaped PRFM, to represents diversity in practical application of PRFM in different clinical conditions (Figure 3). Both PRP specimen (disc and dome shaped) centrifuged with 25 mm CaCl_2_ 1M had PPP of 0/μL.

**Figure 2.**
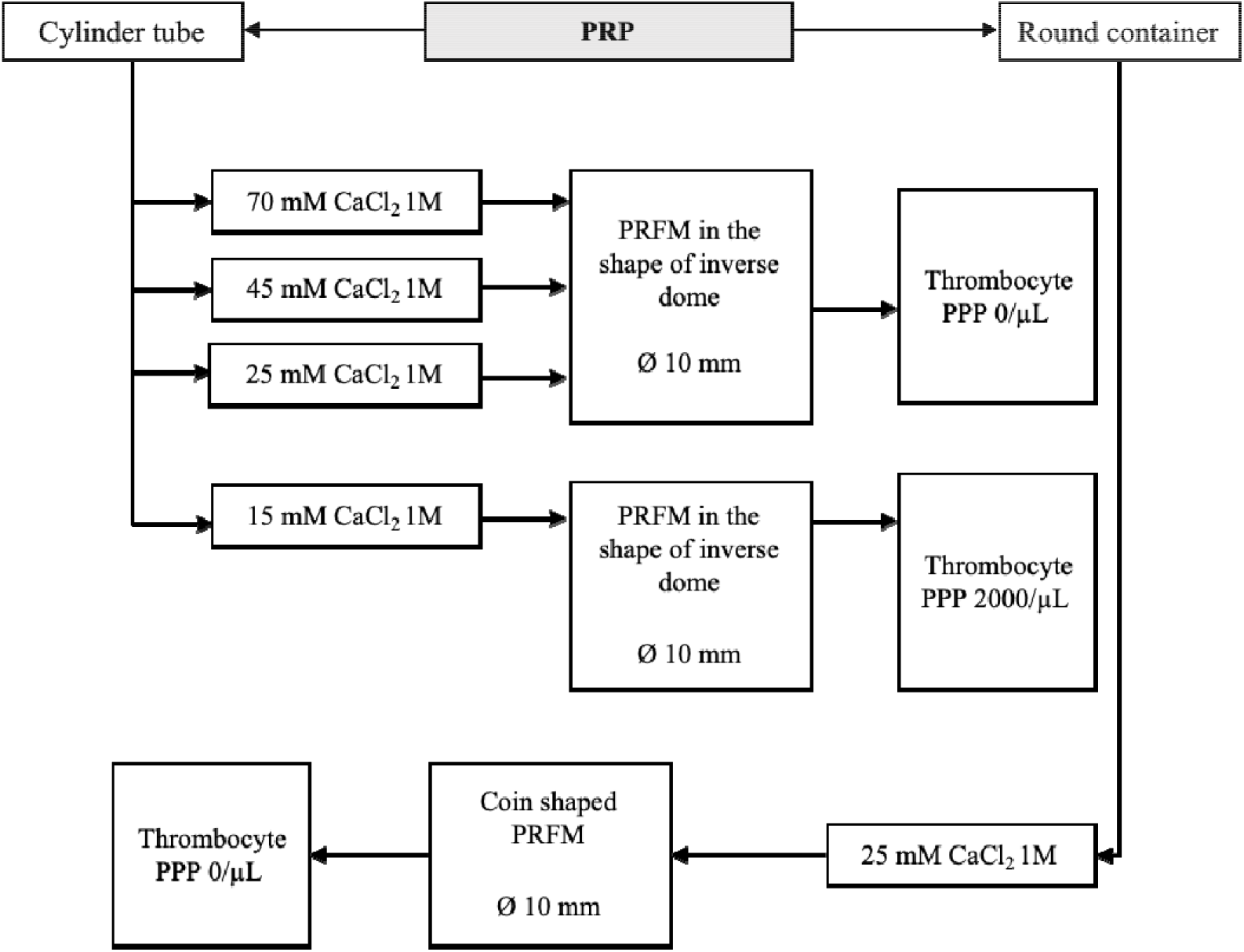
Finding the minimum recommended amount of CaCl_2_ 1M for ideal PRFM. Regardless of the shape, 25 mM of CaCl_2_ 1M is determined to be the minimum concentration needed to produce PRFM with 0/μL of thrombocyte in its PPP.

**Figure 3.**
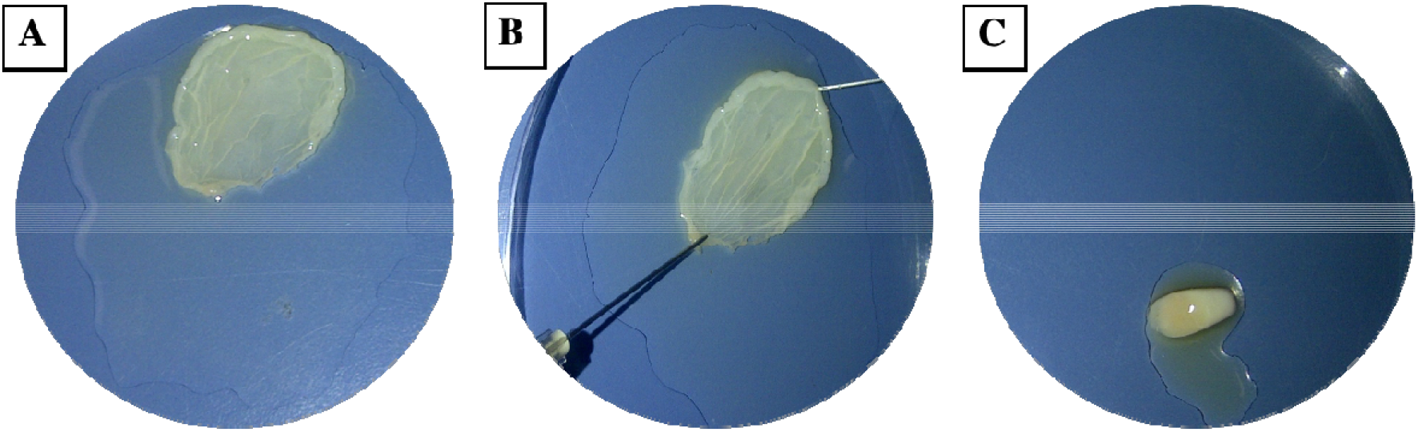
Coin shaped PRFM produced via centrifuging PRP in a round plastic container (A, B) and inverse dome PRFM produced by centrifugation in a cylinder tube (C). The variability in shape produced shows flexibility in PRFM clinical application.

### 3.3. PRFM analysis with SEM

Measurement of fibrin fiber and platelet diameter in PRFM specimens were performed using SEM. PRFM from 5 volunteers was prepared and fixated to produces slices that were thin enough to be analyzed by SEM. It was found that PRFM had platelets scattered among the fibrin fibers (Figure 4). The average diameter size (from three measurements in each PRFM specimens) of the subject’s platelets and fibrin fibers matrix can be seen in the table below (Table 2). PRFM from Subject 1 dried out during preparation and fixation process, rendering it unsuitable for analysis.

**Table 2.** Average fibrin size and platelet diameter in PRFM analyzed through SEM.

| PRFM | Fibrin Fiber Size (nm) | Platelet Diameter (nm) |
| --- | --- | --- |
| Subject 1 | * | * |
| Subject 2 | 446,7 | 670,1 |
| Subject 3 | 403,6 | 638,2 |
| Subject 4 | 360,2 | 625,4 |
| Subject 5 | 428,1 | 638,2 |

\*PRFM dried out during specimen preparation

**Figure 4.**
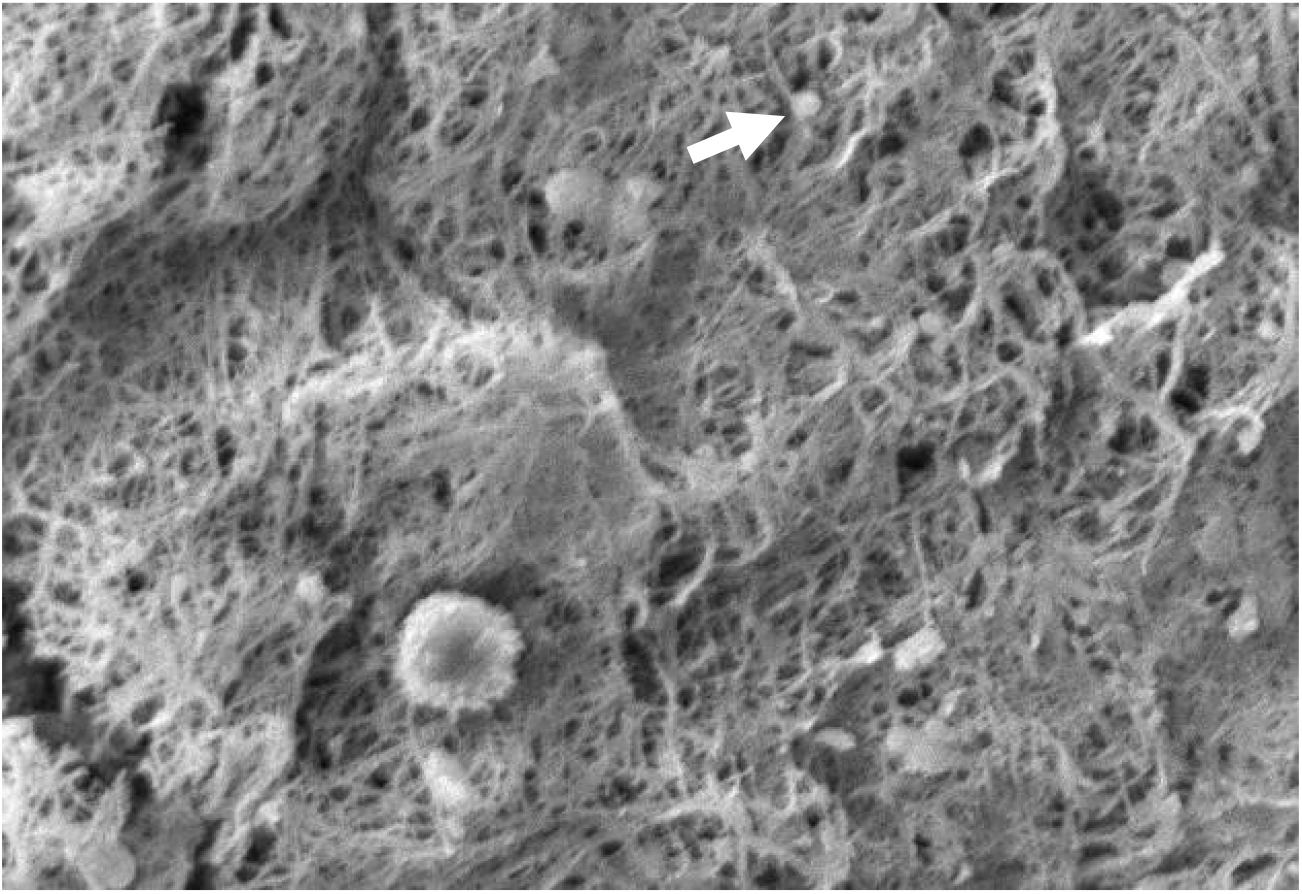
The appearance of fibrin and platelet fibers on PRFM by SEM examination. The white arrows show the platelets between the fibrin fibers matrix. It appears that the fibrin fibers are evenly distributed like a mesh. 1000x magnification.

### 3.4. TGFβ1 in PRFM

The average TGFβ1 rate from PRFM was 37,497 pg/mg, with the highest rate being 42,147 pg/mg and the lowest being 31,849 pg/mg (Table 3). PRFM from Subject 1 dried out during preparation and fixation process, rendering it unsuitable for analysis.

**Table 3.**
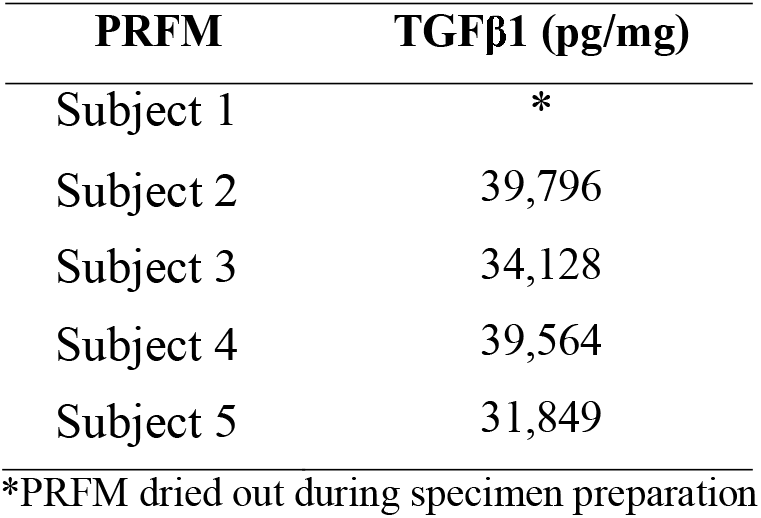
Level of TGFβ1

## 4. Discussion

Currently, several methods and commercial kits are available for the preparation of PRP. Most of this method will produce an end product in liquid or gel form. Due to these mechanical properties, conventional PRP is often impractical in clinical settings that require secure implantation in a specific site or where released growth factors could be washed out during an operation.

The latest development in PRP use for wound healing involves altering its physical property through plasma and platelet stimulation. Alteration of PRP physical properties can be achieved by adding calcium (CaCl_2_) and centrifugation to produce PRFM without the need for additional exogenous thrombin. The addition of CaCl_2_ and centrifugation to PRP will convert fibrinogen to fibrin, and the fibrin cross-links to form a matrix that contains viable platelets (Figure 3). The PRFM preparation process creates a gel-like matrix containing high concentrations of non-activated, functional, intact platelets within a fibrin matrix. These platelets had been proven to release a relatively constant concentration of growth factors over seven days.^9^

The resulting PRFM is a thin sheet with a more robust physical structure than the liquid PRP. The PRFM can replicate a natural wound healing response (i.e., the three-dimensional form of a cross-linked fibrin matrix). This scaffold-like fibrin matrix is essential as a place for platelet adhesion. This scaffolding helps localize platelets and ultimately increases the concentration of growth factors to the desired point or location for tissue regeneration.^10^

This experiment had successfully produced an ideal PRFM through a modified method without using any commercial reagent kit. The ideal PRFM requires that maximal platelets are trapped in the PRFM; the PPP platelet count of 0/mL can prove this. This experiment also found the minimum amount of CaCl_2_ and the centrifuge settings needed to obtain the ideal PRFM. To achieve ideal PRFM, PRP obtained was mixed with 25 mM CaCl_2_ 1M, then centrifuged again at a speed of 2264 G for 25 minutes at room temperature. The centrifugation will result in two layers, the PPP and PRFM. The platelet level in PPP was 0/μL; thus, it can be assumed that this method produces ideal PRFM with all the platelets from PRP adhering to the matrix fibrin. Commercial kits usually include CaCl_2_ solutions in their package, though most choose not to disclose the amount or concentration of CaCl_2_ used in their set.

The PRFM protocol for this study is based on a previous experiment by O’Connell, which breakdowns the creation of PRFM from whole blood. O’Connell created PRFM by inserting PRP and CaCl_2_ 1M in a Wheaton bottle before starting the centrifugation. In his study, 18 mL of whole blood can create 7–8 mL of PRP, which yields a 35 mm round PRFM membrane.^8^ In our study, 8 mL of whole blood produces 6 mL of PRP, which then yields a 10 mm round PRFM membrane. A suspected vital difference in methodology lies during the PRP creation in which tubes containing a thixotropic polyester separator gel was used. These separator gels contain different properties depending on the PRP kit’s brand (O’Connell used tubes from the Cascade Autologous System, while this study used one from RegenLab). Differences in the separator gel may affect performance in isolating platelets and plasma (containing fibrinogen) from the packed red and white cell fraction.^8,11^

The platelet count results for whole blood, PRP and PPP after the first centrifugation showed that the platelet count in PRP was lower than that of the whole blood (Table 1). In general, the preparation of PRP is followed by volume adjustment by removing the PPP, resulting in a platelet concentration 2.5 to 8 times higher than that found in whole blood.^12^ This study’s median PRP platelet count was 97,800 (52,000-186,000)/mL – up to 50% loss of platelet, which is lower than the typical PRP platelet count.^13^ The low platelet count is probably caused by the trapping of platelets in the RegenKit gel tube or due to erythrocyte deposits (Figure 5). Unfortunately, no literature specifically discusses platelet yield in PRP production. Platelet yield must be considered in assessing the PRP creation methods used, including standardised methods (commercial kits). Ideally, efforts are made to minimise platelets lost since these platelets are a source of growth factors.

**Figure 5.**
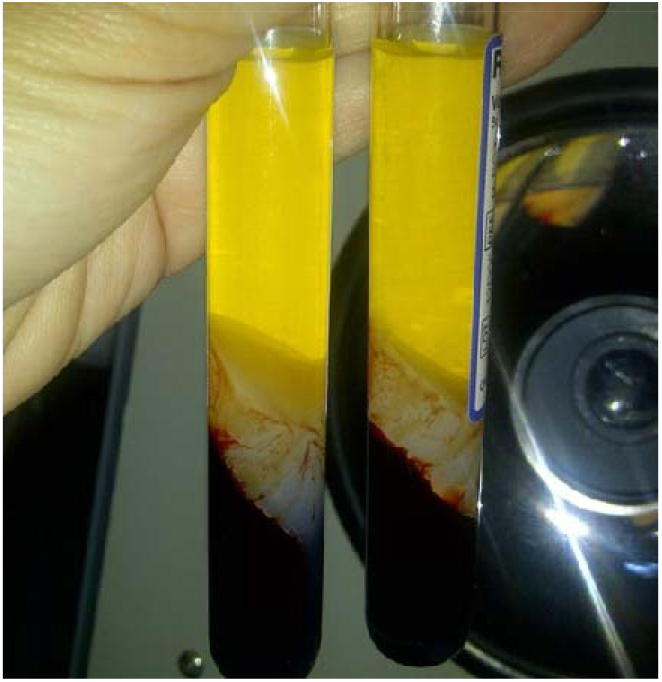
RegenKit tube with separator gel in the middle. Platelet might be trapped in the gel tube thus resulting in lower platelet count in the resulting PRP.

Based on SEM examination, it can be seen that the PRFM obtained has a microscopic fibrin fibre matrix resembling a mesh (Figure 4). All platelets derived from PRP are evenly distributed among the fibrin fibres matrix. Thus, it can be concluded that this PRFM preparation method consistently produces microscopic fibrin fibre matrix with identical platelet distribution present in ideal PRFM.^10^ The resulting PRFM is denser and more flexible, resembling a fascial layer that can be sewn, identical to PRFM preparation resulting from a commercial kit.^5^

Other properties analyzed in the PRFM produced are TGFβ1, a growth factor responsible for controlling and promoting cell growth, proliferation, and differentiation.^14^ Contrary to popular belief, usually, the TGFβ1 level in PRP is higher than the PRFM. The difference in TGFβ1 level can occur due to the activation of platelets by exogenous factors during PRP and PRFM preparation. These exogenous factors can occur during venous blood collection, pipetting, or in the centrifugation process.^15–17^ Activated platelets will release granules along with their contents, including growth factors such as TGFβ1 and PDGF, which will then dissolved in the plasma. During PRFM preparation, the centrifugation process at 1,800 G for 60 minutes will precipitate platelets and the formed fibrin polymer. The dissolved protein will remain in the plasma, resulting in TGFβ1 and the activated platelets not precipitating in PRFM. It is known that both cytokines and growth factors secreted from cells have a short half-life, which means that a higher level of TGFβ1 in PRP does not mean that PRP is better than PRFM.^18^ Comparison of TGFβ1 between PRP and PRFM might not yield any meaningful, practical result, yet it would be interesting to look upon the correlation of the initial amount of TGFβ1 in PRFM and the amount of TGFβ1 released after the PRFM had been administered to a media or wound site.

The weakness of this study is a lack of actual observation and comparison towards actual clinical use of the PRFM produced through the modified method proposed. A follow-up study looking at how the PRFM produced through this method fare compared to PRFM produced by commercial kit would further raise the credibility of using the proposed method as alternative ways of preparing PRFM, especially in limited-resource settings. Nevertheless, the study had proven that this modified method could produce ideal PRFM with somewhat comparable quality to the ones produced using a commercial kit.

## 5. Conclusion

The proposed modified method by mixing PRP with 25 mM CaCl_2_ 1M and centrifuging at a speed of 2264 G for 25 minutes at room temperature can reliably produce ideal PRFM comparable in quality to the commercial kit. Further follow-up study is needed to compare the performance of PRFM produced by the modified method to those produced with commercial kits.

## Data Availability

Data from this study are available upon request

## 6. Acknowledgement

The author would like to thank friends and fellow staff in University of Indonesia who had supported the study.

## Notes

### Competing Interest Statement

The authors have declared no competing interest.

### Funding Statement

No external funding was received for this study

### Author Declarations

This study is conducted in Indonesia and approved by the Ethical Clearance Committee from the Faculty of Medicine, University of Indonesia (202/H2.F1/ETIK/2013).

### Summary of Updates

- Added ORCID ID for all the other authors in the author list - Correction in section 2.1 (the first sentences incorrectly refers to the previous Table 1 which had been omitted from the paper)

